# Childhood Socioeconomic Status and Polygenic Scores for Cognition Have Independent Associations with Cognitive Performance During Middle Childhood

**DOI:** 10.1101/2021.08.26.21262684

**Authors:** S.E. Paul, N.M. Elsayed, R. Bogdan, S.M.C. Colbert, A.S. Hatoum, D.M. Barch

**Author notes:** Correspondence concerning this article should be addressed to Nourhan M Elsayed, address: Somers Family Hall Room 325A, Washington University in St. Louis, One Brookings Drive, St. Louis, MO 63130, USA). These authors contributed equally to this work (S.E. Paul, N.M. Elsayed). These authors share senior authorship (A.S. Hatoum, D.M. Barch).

## Abstract

Childhood cognitive abilities are heritable and influenced by malleable environmental factors such as socioeconomic status (SES). As cognition and SES share genetic architecture, it is critical to understand the extent to which SES is associated with cognition beyond genetic propensity to inform the potential benefit of SES-based interventions. Previous investigations conducted in small samples have suggested that SES is linked with cognitive ability independent of polygenic prediction for educational attainment. Here, we extend this work to a large sample (total n = 4,650) of children (ages 9-10) of genomically-confirmed European ancestry. We find that an SES composite (i.e., family income-to-needs, caregiver education, and neighborhood median income) and a polygenic cognition score composite created using genomic structural equation modeling (COG PGS; Educational Attainment, Intelligence, and Executive Function) are associated with cognitive performance indices (i.e., general ability, executive function, learning/memory, fluid intelligence) that are largely independent of one another. SES x COG PGS interactions are not associated with cognition. These findings provide further evidence for the significant role of modifiable environmental factors in the development of cognitive abilities in youth.

## BACKGROUND

Childhood poverty is robustly associated with general and specific cognitive performance deficits. ^1–4^ Beginning as early as six months old, youth raised with lower household incomes exhibit poorer performance on measures of total IQ ^5^ as well as specific cognitive processes (e.g., executive function, memory).^6–8^ Greater educational attainment among children whose families received supplemental income ^9–11^ and boosts in cognitive performance induced by enriched environments in non-human animal models ^12^ highlight the plausibility that socioeconomic status (SES) may have a causal impact on child cognition and heighten the urgency of addressing the epidemic of childhood poverty. ^13,14^

At the same time, the moderate heritability of cognitive ability ^15^ and SES, as well as their shared genetic architecture (rg=0.65-0.82 in adults) ^16,17^ has been used to argue that cognitive deficits related to childhood poverty may be partially attributable to shared genetic liability. ^18^ Shared genetic liability may arise from genetic inheritance that directly influences cognitive ability as well as gene-environment correlations that may be more amenable to environmental intervention.

Disentangling genetic and socioeconomic status associations with cognition is challenging. For instance, twin studies may not be able to detect genetic influences on SES or the influence of SES on cognitive development, as family-level SES is typically shared within a twin pair. ^18^ Another approach is to measure genetic influence using polygenic scores (PGS) that effectively represent genome-wide genetic liability to a particular phenotype on an individual level. ^19^ Few studies have directly explored the unique or interacting contributions of PGS and SES in the prediction of cognitive abilities. Relatively small investigations (Ns ≤ 551) have found that both PGS for educational attainment (a frequent proxy for cognitive ability in genetics studies) and SES independently predict specific aspects of cognition (e.g., memory, working memory). ^20,21^ Larger investigations that more comprehensively assess PGS and cognitive abilities are needed. Further, work in the Adolescent Brain Cognitive Development Study (ABCD) sample has shown that both family income ^22^ and neighborhood disadvantage (median neighborhood income) have unique associations with neurocognitive performance in youth. ^23,24^ Thus, it is critical to measure SES comprehensively including both the familial and neighborhood levels, as both are important predictors of cognitive development.

The present study tested the hypothesis that SES uniquely contributes to variance in four domains of cognitive ability (i.e., general ability, executive function, learning/memory, fluid intelligence) that is separable from genomic influence (polygenic scores [PGS] for cognition). Given mixed evidence that SES may moderate the heritability of cognitive outcomes (e.g., ^25,26^), we also tested an SES x PGS interaction in a sample of 4,650 children (ages 9-10) of genomically-confirmed European ancestry. We formed a composite measure of SES from familial income-to-needs, caregiver education, and median neighborhood income to increase comprehensiveness. Likewise, we employed genomic Structural Equation Modeling (GSEM ^27^) to generate a one-factor multivariate GWAS of cognition from three sets of summary statistics (Educational Attainment, N=766,345 ^28^; Executive Function, N=427,037 ^29^; and IQ, N=269,867 ^30^), from which we computed a single PGS for cognitive ability. Linear mixed effect models (to account for data clustered by research site) were conducted. Model parameters were estimated with K-fold cross-validation, and model generalizability was tested in a non-overlapping test set.

## METHODS

### Statement on ethical regulations

Parents/caregivers provided written informed consent, and children verbal assent, to a research protocol approved by the central institutional review board at the University of California at San Diego for 20/21 data collection sites across the United States (https://abcdstudy.org/sites/abcd-sites.html) at by the Washington University IRB for the Washington University site.

### Participants

Data came from 11,875 children (mean ± SD age = 9.91±0.62 years; 47.85% girls; 74.13% white) who completed the baseline assessment of the ongoing longitudinal Adolescent Brain Cognitive Development (ABCD) Study (release 3.0.1; https://abcdstudy.org/). ^45^ The study includes multiple sibling and twin pairs and triplets as part of its family-based design.^46^ Primary analyses will be restricted to genomically-confirmed participants of European ancestry with available genetic data (n = 4,650).

### Measures

#### Demographic Measures

##### Child and parent demographics

Child age was self-reported and measured in months. Child sex was a caregiver-reported dichotomous variable. Caregivers reported on their educational attainment within the phenX Toolkit. ^47^

##### Familial Income

Familial income was estimated using the income-to-needs ratio (INR) from the baseline assessment, consistent with methods described elsewhere. ^48^ Briefly, binned gross household income and the number of household members were reported by caregivers. Binned income levels were adjusted to the median for each bin and divided by the 2017 federal poverty threshold for the given household size to derive the INR. An INR of 100% indicates that a family is at the federal poverty line.

##### Neighborhood poverty

The child’s primary residential address at baseline was geocoded by the Data Analysis, Informatics and Resource Center of the ABCD Study, and variables from the American Community Survey (5-year estimates from 2011 to 2015) were linked to each individual according to their US census tract. Median family income in the area deprivation index was selected as a proxy for total neighborhood poverty.

##### Composite SES Measure

The INR, parental education, and neighborhood poverty were all scaled and averaged to form a composite measure of SES.

#### Cognitive measures

##### Cognitive Ability

Three principal components previously derived in the ABCD sample representing general ability, executive function, and learning/memory were used to index cognitive ability. ^49^ Briefly, a Bayesian Probabilistic Principal Component Analysis (BBPCA) was applied to cognitive tasks from the NIH Toolbox cognition battery, which assesses executive function, attention, processing speed, working memory, episodic memory, and language; the Rey Auditory Verbal Learning Test, which measures auditory learning, memory, and recognition; and the Little Man Task, which assesses visuospatial processing. ^50^ BPPCA component weights for each participant were made available with the ABCD curated data release 2.0.1. The Matrix Reasoning subtest of the Wechsler Intelligence Scale for Children-Fifth Edition (WISC), which broadly indexes fluid reasoning and cognitive flexibility important for life function ^51,52^ was included as an additional measure of executive function.

###### Polygenic Scores (PGS)

Summary statistics from the most well-powered, publicly available genome-wide association studies (GWAS) of three cognitive phenotypes (Educational Attainment, N=766,345 ^28^; Executive Function, N=427,037 ^29^; and IQ, N=269,867 ^30^) were used to generate a one-factor multivariate GWAS using genomic Structural Equation Modeling (GSEM; **see supplement for more information**). The resulting summary statistics were used to generate PGS in the European ancestry subsample of ABCD (n=4,650). PGS were computed using PRS-CS, ^53^ a Bayesian approach that incorporates all SNPs (i.e., no p-value thresholding) and utilizes an external linkage disequilibrium (LD) reference panel to account for correlations between SNPs. The “auto” function within the PRS-CS software package was used to compute PGS (see **Supplement** for further details).

###### Genotyping, Quality Control, and Imputation

The Rutgers University Cell and DNA repository genotyped saliva samples on the Smokescreen array. Genotyped calls were aligned to GRCh37 (hg19). The genetic data underwent typical quality control procedures following the Ricopili pipeline. ^54^ Analyses were restricted to individuals of genetically confirmed European ancestry, to match the ancestry makeup of the discovery GWAS. Further details are provided in the **Supplement.**

### Analytical Methods

Linear mixed effects models were used to examine the relationship between SES, PGS, and cognition. Each of the four cognition outcomes were examined independently, with PGS and the SES composite included as the primary predictors of interest. All models covaried for sex, age, and the first 10 ancestrally informative principal components (PCs) and included research site as a random effect. Additional models examined whether SES moderated effects of PGS on each cognitive outcome, further covarying for all covariate-by-SES and covariate-by-PGS interactions. ^55^ Using a regression approach to examine GxE allows for cross-validation (see below), mixed models, and computation of the proportion of variance explained in the outcome. Follow-up analyses to our conservative approach using an SES composite were conducted with each SES measure independently. Similarly, supplementary analyses were conducted examining each PGS (i.e., educational attainment, intelligence, executive function) independently (**Supplement**).

#### Potential for Inflation

Additional supplementary analyses were conducted to assess whether the inclusion of a heritable environmental variable (SES) correlated with COG PGS resulted in biased estimates by conditioning a collider (**Supplement**).

#### Multiple Testing Correction

The false discovery rate (FDR) ^56^ was used to correct for multiple testing. Specifically, in primary analyses examining the main effects of the cognition PGS and SES composite on each of four cognitive outcomes, all eight p-values were subjected to FDR correction simultaneously.

#### Cross validation

Cross-validation procedures were used to enhance the robustness and generalizability of our findings. Data were initially partitioned into a training sample for model building and a test sample for validation, accounting for familial structure. Specifically, all singletons were including in the training set and participants with sibling(s) in the study were all included in the test set, to avoid dependencies across the training and test sets. All continuous and ordinal variables were scaled to have a mean of 0 and standard deviation of 1 separately in the training and test samples.

The training set was split randomly into four folds, keeping participants in the same site in the same fold (stratified cross-fold validation). The linear mixed effects models were conducted in the training set, and the parameter estimates from the fold with the lowest Root Mean Square Error (RMSE) were reported. The models were applied to the test set using the predict() function, and the resulting RMSE value was compared to that from the training set. A substantially greater RMSE value in the test set compared to the training set would indicate that the model overfitted the data and does not generalize well out-of-sample.

Further, in order to examine whether the beta estimates for predictors of interest (i.e., SES composite and cognition PGS) predicted well out of sample, scores on these variables in the test sample were multiplied by the beta estimates for these variables generated from the primary multiple regressions. These two variables were then summed and correlated with the relevant neurocognitive outcome to assess whether the predicted estimates were correlated with measured outcomes.

## RESULTS

Our sample of 4,650 children of genetically confirmed European ancestry was divided into a training sample (2,985 singletons (mean [SD] age = 118.6 [7.39] months; 46% female) and test sample (1,665 non-singletons (mean [SD] age = 120.4 [7.65] months; 48% female). The training set was significantly younger, had higher levels of caregiver education and income-to-needs, and exhibited higher scores on General Ability, Learning/Memory, and Matrix Reasoning and lower levels of Executive Function, relative to the test set. See **Supplemental Table 1** for more descriptive information of the sample.

### Association between SES composite and Cognition PGS and Cognition

To determine whether both the SES composite and Cognition PGS (COG PGS) are associated with cognitive ability, we examined the relationship between the SES composite and the COG PGS and each of the measures of cognition first using separate linear mixed effects models in the training set and then including both predictors in each model simultaneously. Each of the four cognition outcomes were examined independently. All models covaried for sex and age and included research site as a random effect. For models with COG PGS, we also controlled the first 10 ancestrally informative principal components (PCs). Models including covariates and COG PGS explained between eight and 21 percent of the variance in cognitive abilities and similarly, models including covariates and the SES composite accounted for between six and 18 percent of the variance in cognition (**Table 1**). Of this variance, COG PGS explained between one and eight percent of the unique variance in cognition, whereas the SES composite explained between one and five percent of the variance in cognition (**Table 1**). The SES composite and COG PGS both explained the most variance in general ability relative to the other cognitive domains (**Figure 1, Table 1**).

**Table 1.** Proportion of Variance Accounted for by COG PGS and SES Composite.

| Model | *b* | 95% CI | *p* | R<sup>2</sup>c |
| --- | --- | --- | --- | --- |
| **General Ability** |  |  |  |  |
| Only covariates | -- | -- | -- | 0.13 |
| Covariates + PGS | 0.30 | 0.26, 0.33 | 8.5e-61 | 0.21 |
| Covariates + SES | 0.26 | 0.21, 0.29 | 8.5e-40 | 0.18 |
| Covariates + PGS + SES | PGS = 0.26<br>SES = 0.20 | PGS = 0.22, 0.29<br>SES = 0.16, 0.23 | 2.4e-46<br>1.6e-25 | 0.24 |
| **Executive Function** |  |  |  |  |
| Only covariates | -- | -- | -- | 0.09 |
| Covariates + PGS | 0.07 | 0.03, 0.11 | 2.0e-04 | 0.09 |
| Covariates + SES | 0.08 | 0.04, 0.12 | 8.7e-05 | 0.09 |
| Covariates + PGS + SES | PGS = 0.06<br>SES = 0.08 | PGS = 0.02, 0.10<br>SES = 0.03, 0.11 | 3.0e-03<br>1.4e-03 | 0.09 |
| **Learning and Memory** |  |  |  |  |
| Only covariates | -- | -- | -- | 0.05 |
| Covariates + PGS | 0.17 | 0.13, 0.21 | 4.2e-18 | 0.08 |
| Covariates + SES | 0.12 | 0.08, 0.16 | 7.4e-09 | 0.07 |
| Covariates + PGS + SES | PGS = 0.15<br>SES = 0.08 | PGS = 0.11, 0.19<br>SES = 0.04, 0.13 | 2.4e-14<br>4.9e-05 | 0.09 |
| **Matrix Reasoning** |  |  |  |  |
| Only covariates | -- | -- | -- | 0.03 |
| Covariates + PGS | 0.22 | 0.18, 0.26 | 1.5e-25 | 0.08 |
| Covariates + SES | 0.17 | 0.13, 0.21 | 1.4e-15 | 0.06 |
| Covariates + PGS + SES | PGS = 0.19<br>SES = 0.13 | PGS = 0.15, 0.24<br>SES = 0.08, 0.17 | 1.7e-19<br>5.7e-09 | 0.10 |

*Note.* R<sup>2</sup>c = conditional R<sup>2</sup>, or the proportion of variance explained by the fixed and random effects.
Betas (*b*) and *p*-values (*p*) represent the beta weights and *p*-values for the Cognitive PGS or SES composite in each respective model.

**Figure 1.**
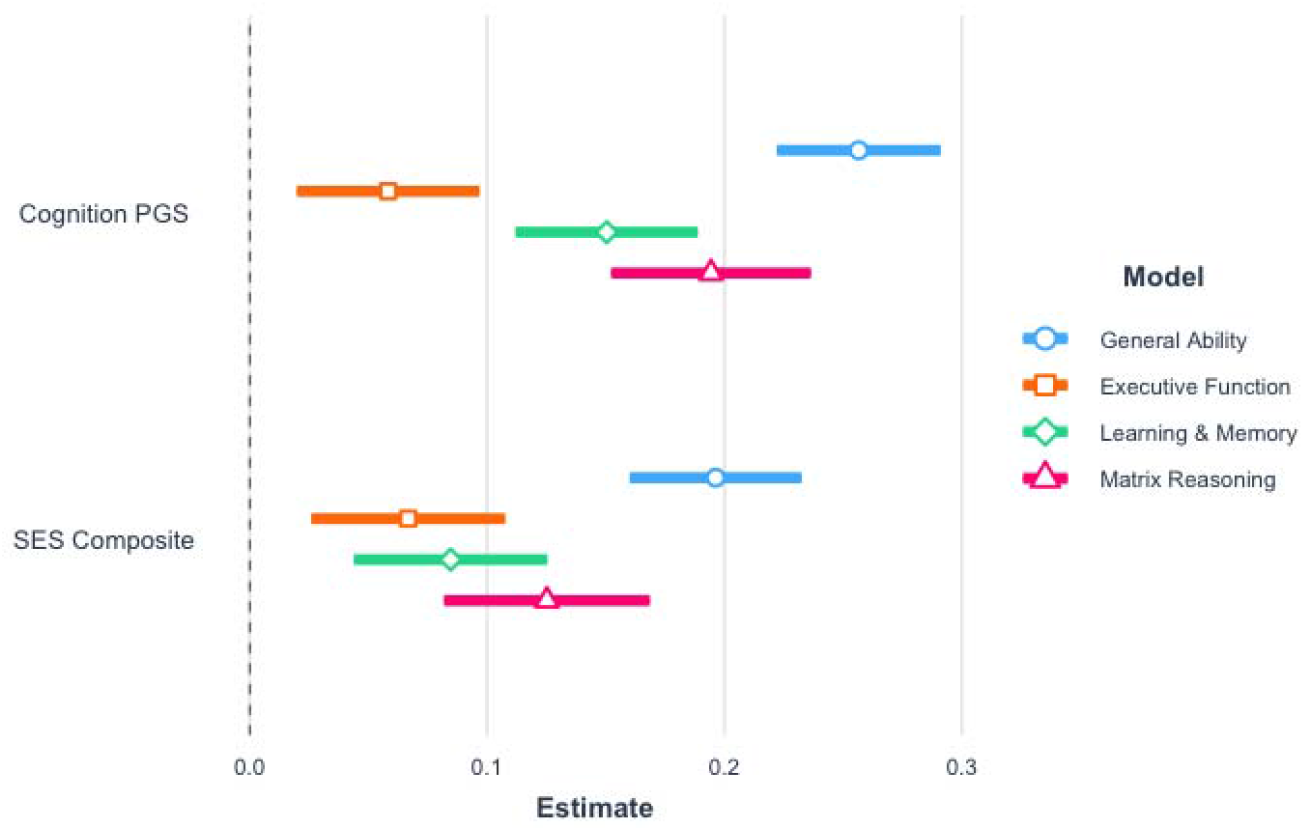
Betas and 95% Confidence Intervals of Cog PGS and SES Composite for Each Cognitive Domain.

To determine whether both COG PGS and the SES composite contributed unique variance above and beyond the other, we then examined their simultaneous contribution to cognition. We found that across all cognitive domains, both COG PGS and the SES composite explained unique variance in cognition (**Table 1**). The inclusion of both predictors simultaneously did attenuate the associations of COG PGS and the SES composite to each cognitive domain assessed; across models, associations were attenuated by an average of 13.64% and 27.8%, respectively (**Table 1**). Individual constituents of the SES composite and COG PGS showed patterns of association with each domain of cognition that were largely consistent with their composites (**Supplement; Supplemental Tables 3-6, Supplemental Figures 3-4).**

### Cross-Validated Models with PGS and SES composite

The linear mixed effects models were conducted in the training set, and the parameter estimates from the fold with the lowest Root Mean Square Error (RMSE) were reported. Cross validation of the above models supported the generalizability of the results to the test sample supported the results of the above analyses; that the COG PGS and the SES composite were each significantly associated with all four measures of cognitive ability, when accounting for covariates and one another (RMSEs: 0.862 - 0.974, **Table 2**).

**Table 2.**
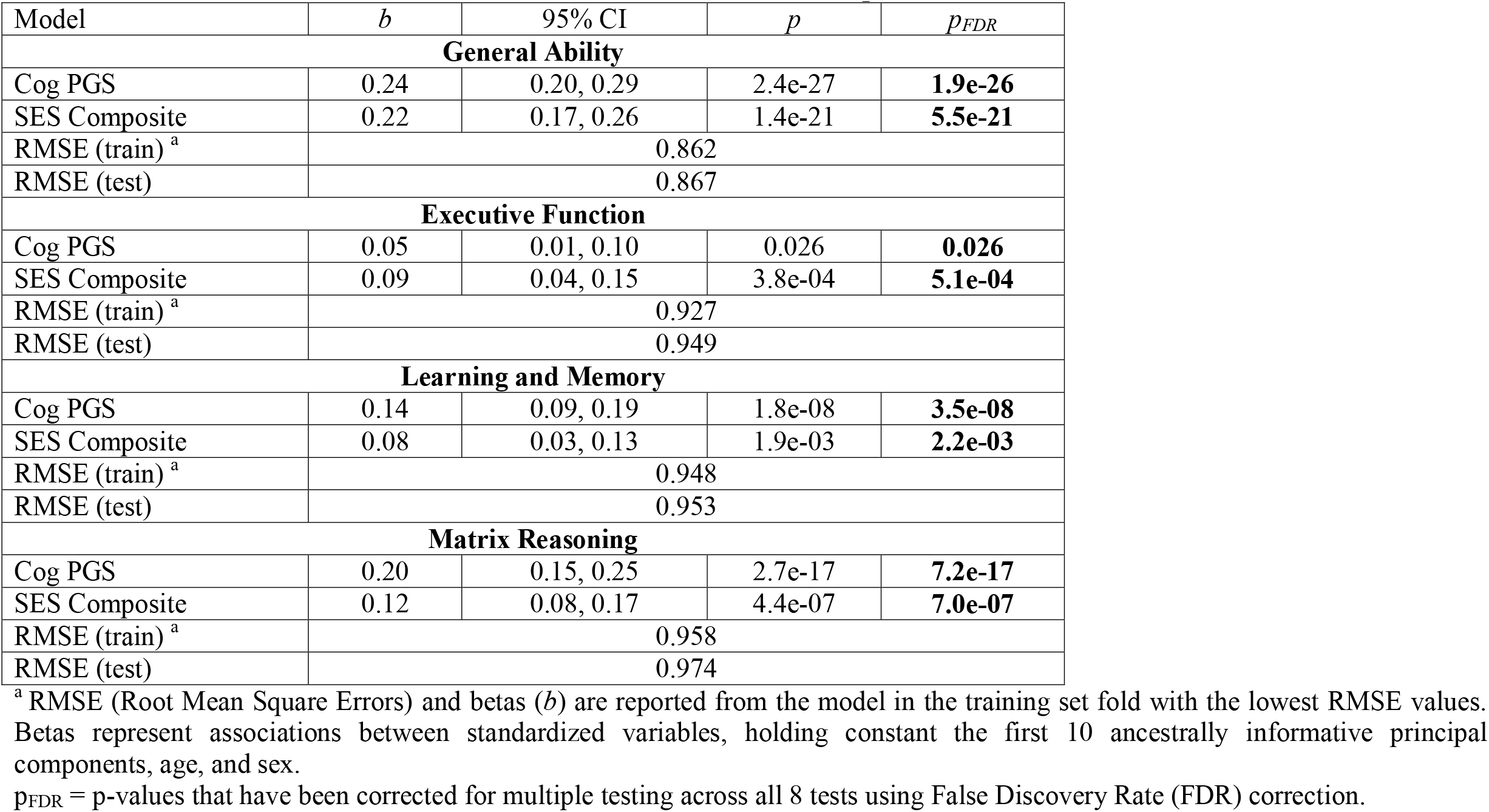
Cross-Validated Results of Model with COG PGS and SES Composite as Simultaneous Predictors.

| Model | *b* | 95% CI | *p* | *p*<sub>FDR</sub> |
| --- | --- | --- | --- | --- |
| General Ability |  |  |  |  |
| Cog PGS | 0.24 | 0.20, 0.29 | 2.4e-27 | **1.9e-26** |
| SES Composite | 0.22 | 0.17, 0.26 | 1.4e-21 | **5.5e-21** |
| RMSE (train)<sup>a</sup> | 0.862 |  |  |  |
| RMSE (test) | 0.867 |  |  |  |
| Executive Function |  |  |  |  |
| Cog PGS | 0.05 | 0.01, 0.10 | 0.026 | **0.026** |
| SES Composite | 0.09 | 0.04, 0.15 | 3.8e-04 | **5.1e-04** |
| RMSE (train)<sup>a</sup> | 0.927 |  |  |  |
| RMSE (test) | 0.949 |  |  |  |
| Learning and Memory |  |  |  |  |
| Cog PGS | 0.14 | 0.09, 0.19 | 1.8e-08 | **3.5e-08** |
| SES Composite | 0.08 | 0.03, 0.13 | 1.9e-03 | **2.2e-03** |
| RMSE (train)<sup>a</sup> | 0.948 |  |  |  |
| RMSE (test) | 0.953 |  |  |  |
| Matrix Reasoning |  |  |  |  |
| Cog PGS | 0.20 | 0.15, 0.25 | 2.7e-17 | **7.2e-17** |
| SES Composite | 0.12 | 0.08, 0.17 | 4.4e-07 | **7.0e-07** |
| RMSE (train)<sup>a</sup> | 0.958 |  |  |  |
| RMSE (test) | 0.974 |  |  |  |

<sup>a</sup> RMSE (Root Mean Square Errors) and betas (*b*) are reported from the model in the training set fold with the lowest RMSE values. Betas represent associations between standardized variables, holding constant the first 10 ancestrally informative principal components, age, and sex.
*p<sub>FDR</sub>* = *p*-values that have been corrected for multiple testing across all 8 tests using False Discovery Rate (FDR) correction.

### Interactions of Cognition PGS and SES on Cognition

The COG PGS x SES interaction was not significantly associated with cognitive abilities (**Table 3**), indicating that SES does not moderate the heritability of cognitive outcomes.

**Table 3.**
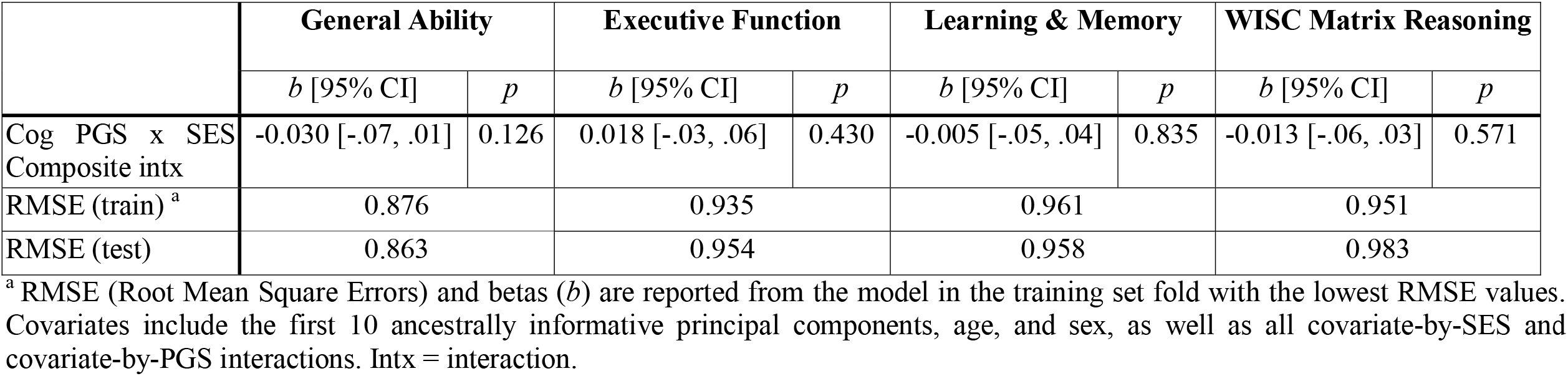
Cross-Validated Results: Models with Interactions.

## DISCUSSION

Here, we demonstrate that socioeconomic status (SES) and genetic (COG PGS) factors are independently associated with multiple indicators of cognitive ability in middle childhood, with no evidence that SES moderates the influence of COG PGS. More specifically, SES and COG PGS were both positively related to General Ability, Executive Function, Learning/Memory, and Matrix Reasoning, and in all cases SES and COG PGS had similar effect sizes with overlapping confidence intervals. These results extend past work showing additive effects of genes and environmental factors (i.e., a lack of dependency)^20,21^ together suggesting that intervention in one aspect (i.e., SES) does not depend on predisposition to higher cognitive abilities.

Broadly, this study extends these previous findings to a larger sample using a more comprehensive measure of genetic influences on cognitive ability and wider range of cognitive outcomes and suggests that public policies that reduce existing socioeconomic disparities such as government-funded supplemental income and universal living wage may enhance cognitive outcomes that augur longer term physical, mental, and financial benefits. As such, programs such as the Advance Child Tax Credit are a likely good first step in supporting the cognitive and psychosocial development of youth.

Generally, both COG PGS and SES were most strongly related to General Ability and least strongly related to Executive Function, as indicated by non-overlapping confidence intervals on the beta weights for these predictors. This may indicate that both SES and PGS are most strongly related to broad cognitive factors that influence of range of function versus more focused cognitive abilities, though it should be noted that the PGS, which itself indexes genomic propensity to broad cognitive ability, would be expected to associated more strongly with broad vs. specific cognitive abilities. Notably, however, prior literature has reported that prefrontal region-mediated cognitive abilities such as executive function are among the most strongly related to SES, although to our knowledge no other studies simultaneously examined both general ability and executive function.^31^ It is also likely however, that our measures of executive function differs slightly from those previously examined.^31^ It is also important to note that each of the individual indicators of SES (INR, caregiver education, neighborhood income) continued to predict cognitive ability with the COG PGS in the same model, and that several SES indicators each accounted for unique variance in cognitive function (e.g., both INR and caregiver education accounted for independent variance in General Ability and Matrix Reasoning, and both neighborhood income and caregiver education accounted for unique variance in Executive Function). These findings are consistent with previous work in this sample, and also extend it by controlling for the influence of COG PGS and by incorporating a broader assessment of SES. ^24^ Analogously, each of the individual PGS also accounted for unique variance in cognition with the SES composite in the models, with evidence that individual PGS each accounted for unique variance in cognitive function when in the same model (e.g., Intelligence and Educational Attainment PGS for General Ability, and all three PGS for Matrix Reasoning). Such findings indicate that while the composite scores for SES and PGS are highly useful and efficient ways for assessing unique environmental versus genetic associations with cognition, further examination of the individual metrics may yield important information about shared versus unique mechanisms and pathways.

The lack of evidence for an interaction between SES and Cognition PGS on cognitive outcomes in the present sample contrasts with earlier findings from twin studies conducted in the United States that show that the influence of additive genetic factors on cognitive ability increases with higher levels of SES, ^26,32,33^ and with one study in the United Kingdom in which polygenic influences on cognitive ability were amplified in the context of more socioeconomic disadvantage. ^34^ However, our findings mirror those primarily from non-U.S. samples that did not find such an interaction, ^25,35,36^ as well as evidence from a more recent, population-level twin study of school-aged youth in the U.S. ^37^ Our results contribute to a complicated and controversial literature by showing that, among a large sample of children from across the U.S., genetic influences on cognition derived from three GWAS of cognitive abilities are not moderated by childhood SES across a range of cognitive functions.

Strengths of this study include the relatively large sample and cross-validation procedures that indicate some level of generalizability of the findings, and the use of composite measures for SES and PGS that enhance power and construct validity. Further, our use of polygenic scores derived from GWAS overcomes some important limitations of twin studies and thus represents a relative strength of this study. In addition to violations of the equal environments assumption^38^, twin studies are unable to isolate or examine interactions with the effect of family SES on cognitive ability, because twins have the same level of SES. Here, by leveraging genome-wide data and measured SES, we were able to estimate the independent effect of SES on cognitive abilities, accounting for genetic influences on cognition that share variance with SES. At the same time, polygenic scores include information only from common genetic variants, and thus, in conjunction with still-limited power of PGS to explain the common variant heritability, are underestimates of the true genetic effect on cognition. Further, rare genetic variants and de novo mutations not incorporated into the PGS may interact with SES, ^34,39^ and our approach was not able to account for this possibility. That we derived our cognition PGS from a multivariate analysis of three GWAS of cognitive abilities, however, improves upon previous analyses of PGS and SES in terms of power and comprehensiveness. However, this study is not without limitations. *First*, the data are cross-sectional, and SES was measured at only one time point. *Second*, the three GWAS we used to compute our PGS were conducted in samples of European ancestry, so we restricted our analyses to individuals also of European ancestry to avoid potential biases.^40^ Given the intersectionality between SES and race alongside typically higher levels of poverty among BIPOC populations, it is problematic that our findings may not be generalizable across all ancestral or racial groups. *Third*, the adjusted R^2^ values from our primary models when including covariates (≤0.24) suggest that large proportions of variance in the cognitive abilities examined are still unexplained. Other influences, including but not limited to caregiver and child stress, mental health, parent behavior, and parent genes for cognition that were not passed to their children but nevertheless shaped their cognitive environment, are also important factors to consider.

*Fourth*, the inclusion of PGS and heritable environmental factors (e.g., SES) in the same regression can create spurious associations. ^41^ In that case, the effect of the environment should increase, while the effect of genetics should decrease. We did not find this to be the case in our models, suggesting that the associations we observed are not spurious. Relatedly, prior work shows that gene x environment interactions may be inflated in the case of gene-environment correlations (i.e., between Cognition PGS and SES ^41–43^). Because we did not detect any interaction between PGS and SES in our models, we expect little inflation of these estimates. *Finally*, despite some differences between the training and test sets (**Supplementary Table 1**), our cross-validation procedures suggested good replication across these samples, enhancing the generalizability of our findings.

Overall, the results of this study provide further evidence that measures of SES are independently associated with cognitive function in children after accounting for genetics, and that the magnitude of the relationship between socioeconomic status and cognitive ability in middle childhood is similar to the effect of genetic influences. These findings highlight the unique importance of a modifiable environmental factor, SES, to childhood cognitive ability and lend further empirical support to those investigating the effects of interventions designed to alleviate childhood poverty, such as direct payments designed to increase family financial stability and enhance child cognitive and psychosocial functioning. ^44^ This work emphasizes the continued need for public policy solutions to the problem of childhood poverty. Future work is needed to examine mechanisms through which both genes and SES may influence cognitive ability (e.g., brain structure, stress, physical and mental health), which may also hold implications for intervention efforts and local and federal policy aimed at reducing child poverty and its sequelae.

## Supporting information

Supplemental methods, results, tables, and figures

## Data Availability

Data belong to the ABCD consortium.

