## Supplemental methods, results, tables, and figures for "Childhood Socioeconomic Status and Polygenic Scores for Cognition Have Independent Associations with Cognitive Performance During Middle Childhood"

**Quality Control.** The Ricopili pipeline was used to conduct the following processing steps. Briefly, SNPs with call rates ≥ 0.95 and minor allele frequency (MAF) ≥ 1% were retained. Individuals with high rates of missingness (>5%) and autosomal heterozygosity deviation (FHET) outside of ± 2 SD were removed. After sample quality control (QC), SNPs were further filtered to call rate ≥ 0.98 and Hardy-Weinberg p-values > 1E-6 (founders only), yielding 372,342 SNPs. Sex checks were conducted with follow-up to reconcile mismatches.

Individuals that passed the first phase of QC were then checked for both known and cryptic relatedness, and Mendelian errors were resolved. Next, using data from unrelated individuals (pi-hat ≤ 0.2) and a linkage disequilibrium (LD) pruned set of common (MAF > 5%) and non-palindromic SNPs (and excluding major histone compatibility complex (MHC) and chromosome 8 inversion region), principal components analysis (PCA) was conducted using the cosmopolitan 1000 Genomes Project phase 3 data in EIGENSTRAT. Only those individuals aligning with non-Hispanic European ancestry were retained, yielding a final sample size of 4,737. After selection, a final ancestrally-informative PCA was conducted, and the first 10 principal components (PCs) were projected from founders to other relatives. Imputation to 1000 Genomes and Haplotype Reference Consortium (HRC) data was performed using strictly QCed SNPs on the Michigan Imputation Server, yielding 39,127,678 SNPs. Dosage data were converted to hard-call genotypes using Plink, and only SNPs with imputation r^2^ scores ≥ 0.3 were carried forward into polygenic scores.

**Genomic Structural Equation Modeling (gSEM).** Using genomic structural equation modeling ^1^, we modeled the genetic correlation matrix of three cognition-related phenotypes using confirmatory factor analysis. We loaded intelligence ^2^, educational attainment ^3^, and executive functioning ^4^ onto a common *Cognition Factor* (**Supplementary Figure 1**). We used the high genetic correlations between these phenotypes (r_g_ = 0.25-0.86, **Supplementary Table 2**) to inform the factor structure and determine that a common factor model was appropriate. Unit variance identification was used to fix the variance of the latent variable to 1.

After assessing the common factor model, we also used gSEM to perform a multivariate GWAS. The common factor GWAS was performed by regressing the Cognition Factor on a SNP (**Supplementary Figure 2**), and then re-running this model for many SNPs. For the purpose of identification in the models with SNP effects, we use intelligence as a marker variable and fix its factor loading to 1. We choose to assign intelligence as the marker variable as it loaded the strongest onto the common factor in the model without SNP effects. The multivariate GWAS function then returns a set of “summary statistics” for the *Cognition Factor* which we then use in subsequent analyses. Effective sample size was estimated for the *Cognition Factor* following preestablished procedures ^5^.

**Multiple Testing Correction.** In supplemental analyses examining the associations between specific SES measures (i.e., income-to-needs, neighborhood median income, and caregiver education), alongside the cognition PGS, and each cognitive outcome, three sets of eight p-values were subjected to FDR correction (i.e., (1 PGS + 1 SES)*4 cognitive outcomes = 8 p-values). Finally, in supplemental analyses in which all three SES measures were entered into linear mixed effects models simultaneously, with the cognition PGS, separately for each outcome, all 16 p-values (i.e., (1PGS + 3 SES)*4 outcomes = 16 p-values) underwent FDR correction. The same process was used for supplemental analyses in which all three PGS were entered simultaneously, alongside the SES composite; **Supplement; Supplemental Tables 3-6**).

**Potential for Inflation.** It is possible that SES is in part a product of PGS for cognition, in which case a multiple regression approach may produce biased estimates by conditioning on a collider ^6^. To assess for this possibility, we conducted analyses in which SES and PGS were entered into separate models, along with covariates, and compared their effect sizes to those when they were entered into the same model simultaneously. An attenuation of the PGS effect alongside an increase in the SES effect would constitute evidence that SES is a collider variable in these models.

**Supplemental Results**

**Associations between Measures of SES and Cognitive and PGS Measures.** We evaluated how each measure of SES was related to cognition when entered into separate analyses, and when entered simultaneously along with the Cognition PGS (COG PGS) and covariates. Correlations between each SES measure and the composite SES measures revealed that across-the-board SES was most strongly associated with General Ability (**Supplemental Table 2**). Consistent with the pattern seen in bivariate correlations, when measures of SES were entered into linear models predicting cognition with COG PGS and covariates, caregiver education exhibited the largest associations with each cognitive ability, with the exception of neighborhood income which showed a larger association with Executive Function (**Supplemental Table 3**). When all three individual SES measures were entered simultaneously into models explaining each measure of cognition, again with COG PGS and covariates, only caregiver education was consistently related to all four cognition measures (*b*s≥0.050, *p*s≤0.030, *p*s_FDR_≤0.044; **Supplemental Table 4**). In the single models, INR was associated with General Ability and WISC Matrix Reasoning (*b*s≥0.060, *p*s≤0.010, *p*s_FDR_≤0.020), and neighborhood income was associated only with Executive Function (*b*=0.057, *p*=0.014, *p*_FDR_=0.024; **Supplemental Table 4**.	As with SES, each PGS (i.e., Educational Attainment, Intelligence, and Executive Function) were entered into separate analyses, along with the SES composite and covariates, to evaluate how these scores contributed to cognition. Each PGS was significantly associated with each of the four measures of cognitive abilities (*b*s≥0.072, *p*s≤6.0e-04, *p*s_FDR_≤6.9e-04), with the exceptions of PGS for Educational Attainment and Intelligence in relation to Executive Function (*b*s≤0.034, *p*s≥0.076; **Supplemental Table 5**). PGS for both Educational Attainment and Intelligence were most strongly associated with General Ability, and PGS for Executive Function was most strongly associated with Learning/Memory (**Supplemental Table 5**). When all three PGS were entered into the same model simultaneously, again along with the SES composite and covariates, PGS for Executive Function was significantly associated with Executive Function, Learning/Memory, and WISC Matrix Reasoning (*b*s≥0.080, *p*s≤4.1e-04, *p*s_FDR_≤7.3e-04), and PGS for Educational Attainment and Intelligence were both associated with General Ability (*b*s≥0.108, *p*s≤1.2e-06, *p*s_FDR_≤3.2e-06) and WISC Matrix Reasoning (*b*s≥0.069, *p*s≤0.012, *p*s_FDR_≤0.017; **Supplemental Table 6**). Educational Attainment PGS and Intelligence PGS were both most strongly associated with General Ability, whereas Executive Function PGS was most strongly related to Learning/Memory (**Supplemental Table 6)**.

**Potential for Inflation.** As presented in **Table 1**, the beta estimates for SES and COG PGS were slightly attenuated by the inclusion of the COG PGS and SES in the models, respectively. Betas for SES in models with covariates but not COG PGS ranged from 0.080 for Executive Function to 0.26 for General Ability. These estimates decreased by 0.01 (for Executive Function) to 0.06 (for General Ability) when COG PGS was included in the models. Betas for COG PGS in models with covariates but not SES ranged from 0.07 for Executive Function to 0.30 for General Ability, and these estimates decreased by 0.01 (for Executive Function) to 0.04 (for General Ability) when SES was included in the models. As the effect of SES did not increase with the addition of PGS, it is unlikely that SES introduced collider bias.

**Supplemental Table 1. Sample Characteristics.**

|  | **Training Sample** | **Test Sample** | **n Complete Data** | **Significantly Different?** |
| --- | --- | --- | --- | --- |
| Age, M (SD) months | 118.6 (7.39) | 120.4 (7.65) | 2,985; 1,665 | p<0.001 |
| Sex, n (%) female | 1,374 (46%) | 802 (48.2%) | 2,985; 1,665 | p=0.17 |
| Caregiver education, M (SD) ^a^ | 17.6 (1.97) | 17.4 (1.94) | 2,985; 1,665 | p=0.002 |
| Income-to-needs, M (SD) ^a^ | 493 (276) | 450 (249) | 2,865; 1,580 | p<0.001 |
| Neighborhood income, M (SD) ^a^ | 748 (440) | 394 (226) | 2,890; 1,561 | p=0.14 |
| Cognitive Outcomes |  |  |  |  |
| General Ability | 0.30 (0.69) | 0.11 (0.70) | 2,815; 1,570 | p<0.001 |
| Executive Function | 0.03 (0.72) | 0.11 (0.68) | 2,815; 1,570 | p<0.001 |
| Learning/Memory | 0.17 (0.67) | 0.13 (0.68) | 2,815; 1,570 | p=0.02 |
| WISC Matrix Reasoning ^b^ | 10.5 (2.9) | 10.2 (2.8) | 2,481; 1,391 | p=0.02 |

^a^ See **Methods** for an explanation of how caregiver education, income-to-needs, and neighborhood income were measured.

^b^ Total Scaled Score

**Supplemental Table 2. Bivariate Correlation Matrix.**

|  | GA | EF | L/M | WISC | EDU  PGS | INT PGS | EF  PGS | COG PGS | INR | EDU | NBHD | SES COMP |
| --- | --- | --- | --- | --- | --- | --- | --- | --- | --- | --- | --- | --- |
| GA | 1 |  |  |  |  |  |  |  |  |  |  |  |
| EF | 0.130 | 1 |  |  |  |  |  |  |  |  |  |  |
| L/M | 0.247 | 0.110 | 1 |  |  |  |  |  |  |  |  |  |
| WISC | 0.32 | 0.111 | 0.279 | 1 |  |  |  |  |  |  |  |  |
| EDU PGS | 0.262 | 0.038 | 0.118 | 0.184 | 1 |  |  |  |  |  |  |  |
| INT PGS | 0.267 | 0.046 | 0.150 | 0.181 | 0.552 | 1 |  |  |  |  |  |  |
| EF PGS | 0.132 | 0.078 | 0.164 | 0.147 | 0.245 | 0.498 | 1 |  |  |  |  |  |
| COG PGS | 0.309 | 0.072 | 0.178 | 0.225 | 0.820 | 0.857 | 0.579 | 1 |  |  |  |  |
| INR | 0.193 | 0.055 | 0.074 | 0.133 | 0.163 | 0.101 | -0.013^a^ | 0.141 | 1 |  |  |  |
| EDU | 0.271 | 0.078 | 0.191 | 0.181 | 0.266 | 0.184 | 0.049 | 0.247 | 0.428 | 1 |  |  |
| NBHD | 0.161 | 0.098 | 0.041 | 0.089 | 0.123 | 0.093 | 0.039 | 0.141 | 0.427 | 0.305 | 1 |  |
| SES COMP | 0.261 | 0.103 | 0.124 | 0.166 | 0.251 | 0.174 | 0.017^a^ | 0.217 | 0.809 | 0.742 | 0.749 | 1 |

PGS = Polygenic Score. GA = General Ability. EF = Executive Function. L/M = Learning/Memory. WISC = WISC Matrix Reasoning. EDU = Educational Attainment. INT = Intelligence. COG = Cognition. INR = Income-to-Needs. NBHD = Neighborhood median income. SES COMP = Socioeconomic Status Composite.

Estimates were derived in the training sample.

^a^ These associations did not reach significance.

**Supplemental Table 3. Individual SES Measures.**

|  | **General**  **Ability** | | | **Executive**  **Function** | | | **Learning &**  **Memory** | | | **WISC Matrix Reasoning** | | |
| --- | --- | --- | --- | --- | --- | --- | --- | --- | --- | --- | --- | --- |
|  | *b* | *p* | *pFDR* | *b* | *p* | *pFDR* | *b* | *p* | *pFDR* | *b* | *p* | *pFDR* |
| Cog PGS | 0.280 | 9.5e-57 | **7.6e-56** | 0.069 | 2.9e-04 | **3.9e-04** | 0.162 | 3.3e-17 | **8.8e-17** | 0.206 | 8.1e-24 | **3.2e-23** |
| INR ^a^ | 0.140 | 5.4e-15 | **1.1e-14** | 0.024 | 0.227 | 0.227 | 0.041 | 0.039 | **0.045** | 0.098 | 1.4e-06 | **2.3e-06** |
| Cog PGS | 0.300 | 2.4e-64 | **1.9e-63** | 0.068 | 3.4e-04 | **5.5e-04** | 0.179 | 5.4e-21 | **1.4e-20** | 0.218 | 1.0e-26 | **4.1e-26** |
| Neighborhood income ^a^ | 0.095 | 2.9e-07 | **5.9e-07** | 0.064 | 1.9e-03 | **2.6e-03** | -0.003 | 0.886 | 0.886 | 0.050 | 0.017 | **0.020** |
| Cog PGS | 0.265 | 2.1e-52 | **1.7e-51** | 0.061 | 1.1e-03 | **1.3e-03** | 0.140 | 1.1e-13 | **1.7e-13** | 0.194 | 1.2e-21 | **3.2e-21** |
| Caregiver education ^a^ | 0.206 | 2.9e-32 | **1.2e-31** | 0.052 | 6.8e-03 | **6.8e-03** | 0.154 | 6.4e-16 | **1.3e-15** | 0.131 | 9.3e-11 | **1.2e-10** |

^a^ The associations between each SES measure, the cognition PGS, and each cognitive ability were examined in separate models (n=12 total).

Betas (b) represent associations between standardized variables, holding constant the first 10 ancestrally informative principal components, age, and sex.

pFDR = p-values that have been corrected for multiple testing. Three sets (one for each SES measure) of eight FDR corrections were conducted. That is, FDR correction was implemented across all p-values in the first 2 rows, then separately for the next 2 rows, and finally for the last 2 rows.

**Supplementary Table 4. Individual SES Measures, Single Model**

|  | **General Ability** | | | **Executive Function** | | | **Learning & Memory** | | | **WISC Matrix Reasoning** | | |
| --- | --- | --- | --- | --- | --- | --- | --- | --- | --- | --- | --- | --- |
|  | *b* | *p* | *pFDR* | *b* | *p* | *pFDR* | *b* | *p* | *pFDR* | *b* | *p* | *pFDR* |
| Cog PGS | 0.236 | 1.4e-35 | **2.3e-34** | 0.048 | 0.018 | **0.029** | 0.133 | 1.4e-11 | **4.6e-11** | 0.185 | 1.6e-18 | **1.3e-17** |
| INR | 0.077 | 3.2e-04 | **7.3e-04** | 0.001 | 0.954 | 0.955 | -0.001 | 0.955 | 0.955 | 0.060 | 0.010 | **0.020** |
| Neighborhood income | 0.035 | 0.103 | 0.126 | 0.057 | 0.014 | **0.024** | -0.044 | 0.048 | 0.064 | -0.007 | 0.762 | 0.870 |
| Caregiver education | 0.169 | 1.7e-15 | **8.9e-15** | 0.050 | 0.030 | **0.044** | 0.165 | 1.4e-13 | **5.4e-13** | 0.109 | 3.1e-06 | **8.2e-06** |

Betas (b) represent associations between standardized variables, holding constant the first 10 ancestrally informative principal components, age, and sex.

pFDR = p-values that have been corrected for multiple testing. All 16 p-values were entered into the same FDR correction.

**Supplemental Table 5. Individual PGS.**

|  | **General Ability** | | | **Executive Function** | | | **Learning & Memory** | | | **WISC Matrix Reasoning** | | |
| --- | --- | --- | --- | --- | --- | --- | --- | --- | --- | --- | --- | --- |
|  | *b* | *p* | *pFDR* | *b* | *p* | *pFDR* | *b* | *p* | *pFDR* | *b* | *p* | *pFDR* |
| EDU PGS ^a^ | 0.204 | 4.6e-29 | **3.7e-28** | 0.028 | 0.156 | 0.156 | 0.092 | 4.4e-06 | **7.0e-06** | 0.136 | 1.4e-10 | **3.9e-10** |
| SES Composite | 0.199 | 4.6e-25 | **1.8e-24** | 0.072 | 6.0e-04 | **6.9e-04** | 0.094 | 9.1e-06 | **1.2e-05** | 0.134 | 1.4e-09 | **2.8e-09** |
| IQ PGS ^a^ | 0.236 | 5.4e-40 | **4.3e-39** | 0.034 | 0.076 | 0.076 | 0.132 | 1.4e-11 | **2.7e-11** | 0.154 | 1.7e-13 | **4.6e-13** |
| SES Composite | 0.214 | 4.3e-30 | **1.7e-29** | 0.074 | 3.3e-04 | **3.8e-04** | 0.096 | 3.3e-06 | **4.4e-06** | 0.143 | 3.4e-11 | **5.4e-11** |
| EF PGS ^a^ | 0.128 | 3.2e-13 | **6.4e-13** | 0.080 | 2.5e-05 | **2.9e-05** | 0.160 | 5.0e-17 | **2.0e-16** | 0.138 | 1.4e-11 | **2.2e-11** |
| SES Composite | 0.253 | 8.7e-40 | **7.0e-39** | 0.078 | 1.2e-04 | **1.2e-04** | 0.116 | 1.2e-08 | **1.6e-08** | 0.168 | 3.5e-15 | **9.3e-15** |

^a^ The associations between each PGS measure, average SES, and each cognitive ability were examined in separate models (n=12 total).

Betas (b) represent associations between standardized variables, holding constant the first 10 ancestrally informative principal components, age, and sex.

pFDR = p-values that have been corrected for multiple testing. Three sets (one for each PGS measure) of eight FDR corrections were conducted. That is, FDR correction was implemented across all p-values in the first 2 rows, then separately for the next 2 rows, and finally for the last 2 rows.

**Supplementary Table 6. Individual PGS Measures, Single Model**

|  | **General Ability** | | | **Executive Function** | | | **Learning & Memory** | | | **WISC Matrix Reasoning** | | |
| --- | --- | --- | --- | --- | --- | --- | --- | --- | --- | --- | --- | --- |
|  | *b* | *p* | *pFDR* | *b* | *p* | *pFDR* | *b* | *p* | *pFDR* | *b* | *p* | *pFDR* |
| EDU PGS | 0.1089 | 1.2e-06 | **3.2e-06** | 0.018 | 0.459 | 0.489 | 0.034 | 0.148 | 0.182 | 0.078 | 1.8e-03 | **2.9e-03** |
| IQ PGS | 0.161 | 8.2e-11 | **6.6e-10** | -0.026 | 0.335 | 0.383 | 0.045 | 0.082 | 0.109 | 0.069 | 0.012 | **0.017** |
| EF PGS | 0.013 | 0.550 | 0.550 | 0.080 | 4.1e-04 | **7.3e-04** | 0.126 | 1.3e-08 | **5.0e-08** | 0.088 | 1.8e-04 | **3.5e-04** |
| Average SES | 0.211 | 1.4e-25 | **2.2e-24** | 0.088 | 5.3e-05 | **1.2e-04** | 0.104 | 8.9e-07 | **2.9e-06** | 0.134 | 1.1e-09 | **5.9e-09** |

Betas (b) represent associations between standardized variables, holding constant the first 10 ancestrally informative principal components, age, and sex.

pFDR = p-values that have been corrected for multiple testing. All 16 p-values were entered into the same FDR correction.

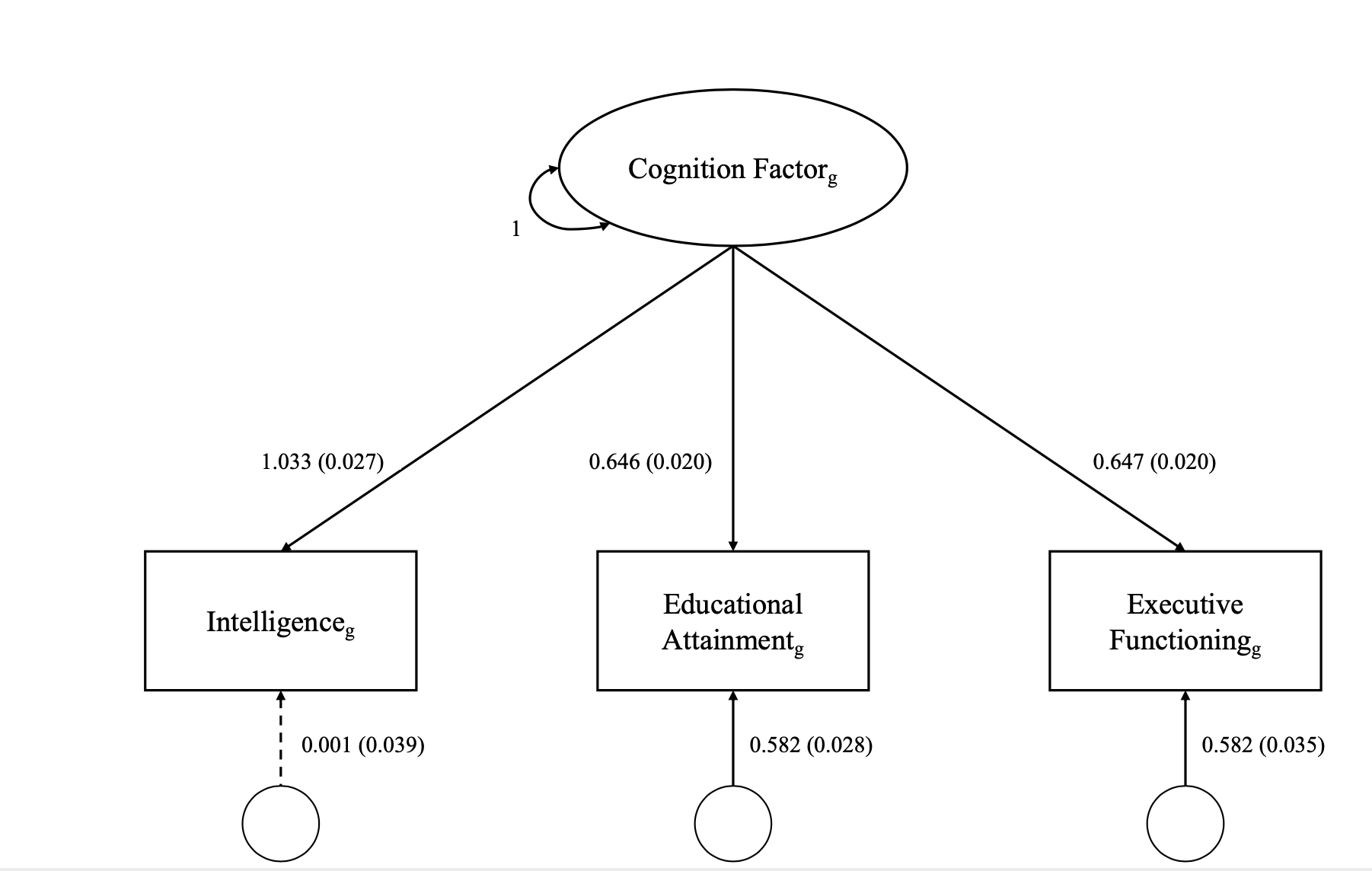

**Supplementary Figure 1.** Results of a loading of intelligence, educational attainment, and executive functioning^4^ onto a common Cognition Factor

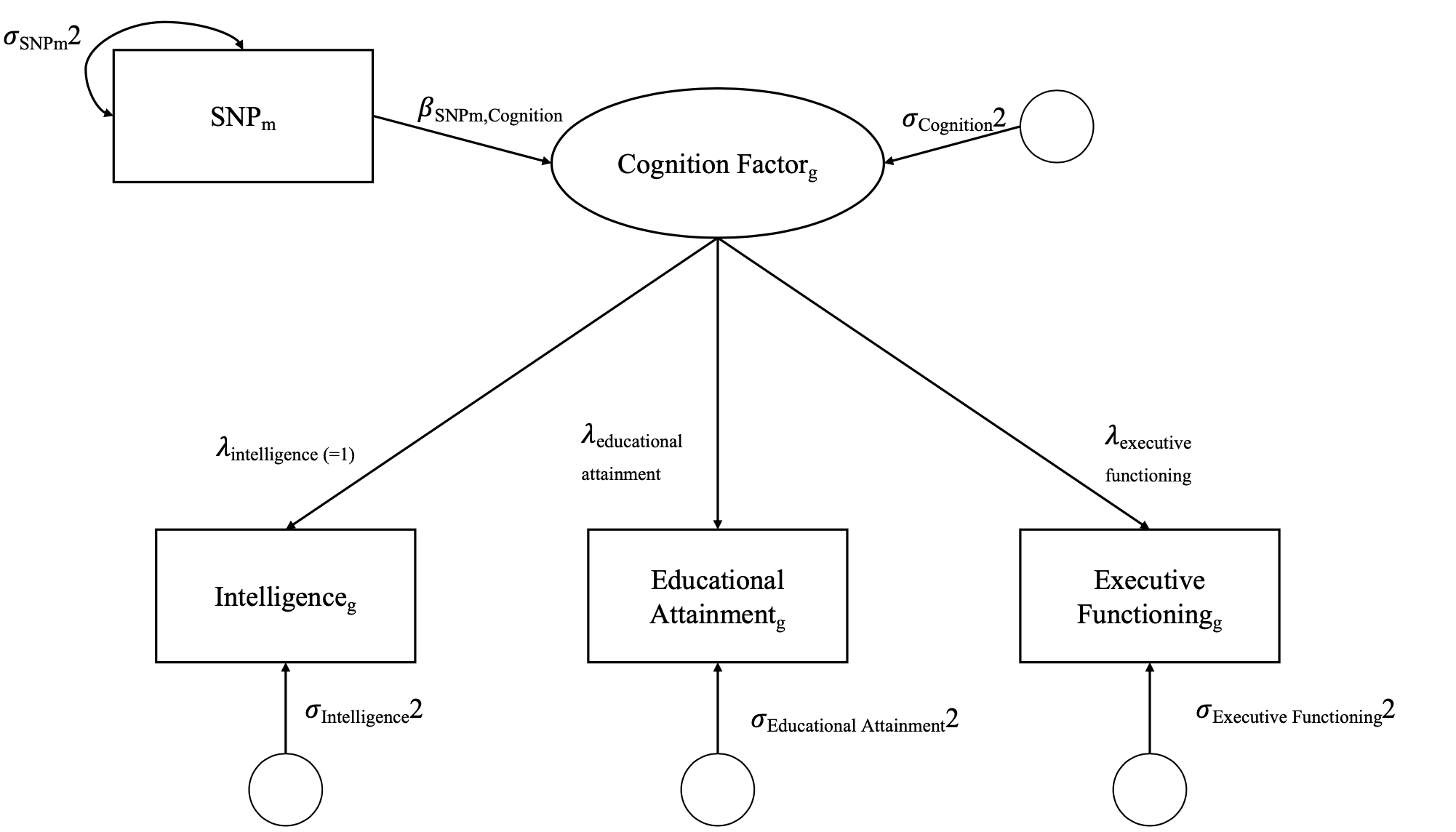

**Supplementary Figure 2.** A common factor GWAS performed by regressing the Cognition Factor on a SNP.

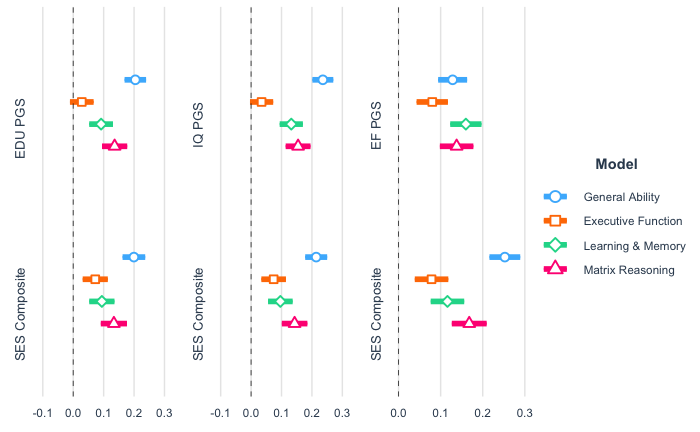

**Supplementary Figure 3.** Betas and 95% CIs of Individual PGS Measures by Domain with SES composite constant

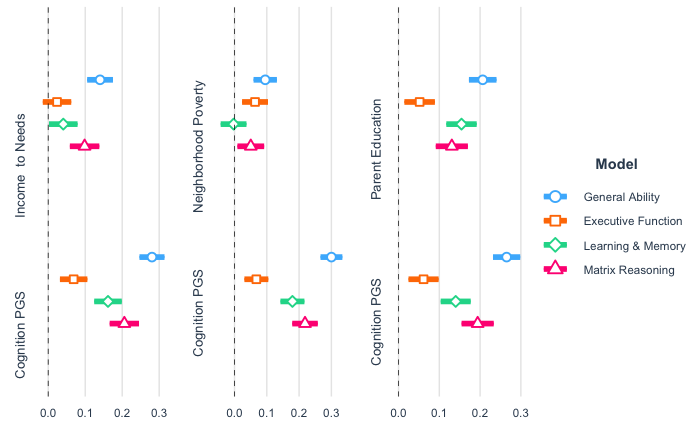

**Supplementary Figure 4.** Betas and 95% CIs of Individual SES Measures by Domain with PGS constant
